# A Multidomain Characterization of Persistent Postural-Perceptual Dizziness Reveals Visual Dependence as Its Main Behavioral Feature

**DOI:** 10.64898/2026.09.22.26363707

**Authors:** Cristina Concetti, Irene Lozzi, Anna Wyss, Salome Häuselmann, Hassen Kerkeni, Tatiana Brémovà-Ertl, Georgios Mantokoudis, Selma Aybek

**Affiliations:** Faculty of Science and Medicine, Department of Movement Science and Neuroscience, University of Fribourg, Switzerland; Department of Neuroscience, Biomedicine and Movement, University of Verona, Italy; Psychosomatic Medicine, Department of Neurology, Bern University Hospital, Switzerland; Dizziness Center, University Clinic for Ear, Nose, and Throat Disorders, Bern University Hospital, Switzerland

**Keywords:** PPPD (Persistent Perceptual Postural Dizziness), FND (Functional Neurological Disorder(s)), Stress markers, Gait, Posture, Vision

## Abstract

**Background:** Persistent postural–perceptual dizziness (PPPD) is a chronic functional dizziness disorder characterized by unsteadiness, hypersensitivity to motion and visually complex environments. Although increasingly investigated, the role of the different factors involved remains unclear. This study aims to comprehensively characterize PPPD across clinical, biological, and behavioral domains.

**Methods:** Data from 18 PPPD patients and 20 age- and sex-matched healthy control volunteers (HC) were analyzed, including clinical and psychological questionnaires, a neuropsychological battery, salivary markers, posturography under different conditions, and gait analysis under single- and dual-task demands.

**Results:** Patients reported greater depressive symptoms, higher perceived stress, and reduced quality of life than HC, but showed no neuropsychological deficits and comparable past psychological trauma and personality traits. Salivary stress marker levels were similar to HC. Gait analysis revealed a slower, cautious gait in patients, which worsened without vision, with no differences in automaticity (stride time variability). Postural assessments showed greater sway and enhanced low-frequency oscillations, exacerbated by absence of visual input in PPPD. Visual reliance for postural control correlated negatively with subjective symptom severity, but not other symptom-related measures.

**Conclusion:** PPPD patients have lower mood and increased perceived stress, with no increase in biological stress markers compared to HC. Cognition, personality, and exposure to past trauma might be less relevant than previously hypothesized. Patients show visual dependence for gait and postural control, likely a maladaptive compensatory mechanism. These findings support interventions targeted at reducing visual dependence and at improving mood, rather than interventions focusing on psychological trauma.

**Summary:**

- **What is already known on this topic** – PPPD (Persistent Perceptual Postural Dizziness) is a functional dizziness disorder characterized by different biopsychosocial factors, often investigated in isolation.
- **What this study adds** – PPPD patients have higher depression and perceived stress scores, lower quality of life, but show no differences in past trauma, personality traits, and biological stress markers, compared to healthy controls. They also show a reliance on vision both for gait and for postural control, with increased low-frequency oscillations in the anteroposterior direction. Visual dependence is likely maintained by its perceived role in symptom reduction.
- **How this study might affect research, practice or policy** - These findings support rehabilitation strategies aimed at reducing reliance on vision and increasing reliance on vestibular inputs, but not interventions focusing on past traumatic experiences.

## Introduction

Persistent postural-perceptual dizziness (PPPD) is a chronic functional neurological disorder (FND), characterized by non-spinning vertigo and subjective postural instability without vestibular or neurological deficits [1–3]. It is one of the most common causes of chronic dizziness[3], with considerable burden for patients[4] and healthcare systems. Patients report feelings of unsteadiness or swaying exacerbated by upright posture and exposure to complex visual stimuli[5].

The control of balance requires the integration of visual, somatosensory, and vestibular inputs. PPPD can emerge after an acute vestibular insult or other stressors that might drive the down-weighting of vestibular inputs and the up-weighting of visual inputs[6]. This response is adaptive when temporary, but maladaptive if maintained chronically, after the resolution of the initial insult[6]. Patients adopt an excessive monitoring of posture that interferes with automatic control, which can in turn further promote stiffened postural strategies and overreliance on visual cues for orientation and balance[7]. This significantly impairs daily activities and quality of life, often co-occurring with anxiety and hypervigilance that exacerbate distress[4,8].

Despite growing recognition of PPPD’s prevalence and impact, its pathophysiological mechanisms remain incompletely understood. Several studies to date have investigated behavioral and motor responses[5,9], psychological factors[10] and brain activity[6].

At the psychological level, PPPD often presents comorbid anxiety and depression[3]. Past psychological traumatic experiences have been proposed as a potential risk factor in PPPD, similarly to other types of FND[2,11], but research on their role in PPPD is scarce[12]. Another proposed risk factor is the personality trait of neuroticism (higher reactivity to stress and tendency to focus on negative experiences[13]). A study on chronic subjective dizziness (a past diagnosis that converged in the new label of PPPD[1]) found higher neuroticism and lower extraversion in patients[14], but this finding awaits confirmation in PPPD. One report shows that PPPD patients also present cognitive difficulties[15], a feature common to some other FND patients[16] and other vestibular disorders[17], highlighting the need to further assess this domain.

At the biological level, alterations in salivary stress markers cortisol (which reflects HPA axis activation[18]), and α-amylase (mediated by the SAM axis[18]), have been found in FNDs [19,20]. To date, stress markers have not been investigated in PPPD, but a relationship between visual dependence for postural control and cortisol reactivity to psychosocial stress has been found in healthy individuals[21].

At the level of motor behavior, the gait of PPPD patients often appears stiff[1]. However, despite this domain being directly relevant to daily life, few previous studies have investigated gait characteristics in PPPD, reporting a lower gait quality[9,22]. More evidence is available on postural control in PPPD[1,23], a direct readout of its symptoms, through posturographic testing or motion sensing, reporting a visual dependence in patients, with unsteadiness increasing with closed eyes[9,24] or moving visual stimuli[23,25]. PPPD patients also reported a higher level of postural instability than objectively measured[26].

Therapeutic advancements for PPPD currently face several challenges, such as disentangling PPPD from similar disorders, addressing its psychological dimensions, and identifying optimal and standardized treatment protocols. Given the multifactorial nature of PPPD, there is a need for integrated investigative approaches to capture the complex interplay of several factors, as highlighted by the biopsychosocial model for FND[2]. Additionally, some of these aspects were studied in diagnostic labels that later converged in the definition of PPPD, but which might not represent the same clinical sample (Phobic Postural Vertigo or PPV, space-motion discomfort or SMD, visual vertigo or VV, chronic subjective dizziness or CSD). Moreover, the similarities and differences between PPPD and the broader category of FND are incompletely characterized[2].

To address these open questions, we have conducted a comprehensive, multimodal investigation, investigating clinical measures of psychological state, life quality, neuropsychological tests, biological stress markers, static posturography, and gait analysis. We hypothesize that 1) PPPD patients will report higher levels of anxiety, depression, stress, past traumatic experiences, and higher levels of neuroticism compared to HC; 2) PPPD patients will show higher levels of salivary stress markers; 3) PPPD patients will show altered gait and postural characteristics with visual dependence.

## Methods

### Participants

40 participants were recruited in this study, of which 20 PPPD patients (18 retained for analysis) and 20 sex- and age-matched HC (all retained for analysis). Patients were recruited from the Dizziness Center and the Psychosomatic Medicine Unit at the Inselspital Bern and diagnosed by a neurologist according to the International Classification of Diseases, 11th revision (ICD-11). HCs were recruited via flyers and word-of-mouth. Inclusion criteria were: aged above 16 years; willing to participate in the study; capable of judgement. Exclusion criteria were: severe comorbid psychiatric disorders, neurological conditions (e.g. epilepsy), history of drug or alcohol abuse, past brain surgery; inability to follow the study protocol; contraindications for MRI; pregnancy and breastfeeding. The methods and results pertaining to MRI neuroimaging are described in another paper (Concetti et al., *in prep*).

The study was approved by the Ethical Committee of the Canton Bern (SNCTP000004529, 2020-02283), registered on ClinicalTrials.gov (NCT05086380), and conducted according to the principles of the Declaration of Helsinki. Participants provided written informed consent and were reimbursed for travel expenses. Data was collected during two study visits.

### Demographical Characteristics, Clinical and Personality Questionnaires

Demographic information, such as age, sex, smoking and medication use was collected. All participants underwent a comprehensive clinical and self-report evaluation. Depressive symptomatology was quantified using the BDI-I, while anxiety was assessed through STAI. To evaluate health-related quality of life, participants completed both the SF-36 and the NQoL. Perceived stress was measured via the PSS. Personality characteristics were captured through the Big5T. Finally, exposure to and impact of adverse life experiences were assessed using the TEC and CTQ. To evaluate vestibular-related symptomatology and its impact on daily activities, participants completed the NVI and DHI. Participants additionally assessed the severity of their symptoms on a visual-analog scale ranging from 0 to 100 (0 = perfect health, 100 = the most severe symptoms, Subjective Symptom Severity score, SSS). Clinician-rated symptom severity was assessed with the CGI. (Full names of questionnaires are provided in the **Abbreviation List**, while references are provided in **Supplementary Materials**.)

### Neuropsychological Assessments

Cognitive functioning was assessed with a neuropsychological battery testing attention, executive function, working memory, and spatial memory. The TAP was administered to probe both phasic alertness and working memory capacities. Executive control was examined through the Stroop Test. Visuospatial short-term and working memory were measured using the BTT, whereas verbal working memory was assessed with Digit Span. (Full names of tests are provided in the **Abbreviation List**, while references are provided in **Supplementary Methods**.)

### Salivary Stress Markers

Salivary cortisol and α-amylase were sampled from participants using cotton swabs (Salivette CollectionDevices, Sarstedt) held in the mouth for ~1 min per sample. Samples for the cortisol analysis were collected at home by participants (instructions are detailed in **Supplementary Methods**). Salivary samples for α-amylase analysis were collected immediately before and after MRI imaging on site. All saliva samples were centrifuged at room temperature for 10 min at 3500 rpm and subsequently stored at −20°C. Salivary cortisol and α-amylase concentrations were determined using commercial enzyme immunoassays (Salimetrics, High Sensitivity Salivary Cortisol Enzyme Immunoassay kit, 1-3002; Salimetrics, Salivary Alpha-Amylase Kinetic Enzyme Assay Kit, 1-1902), following the manufacturer’s protocols.

### Gait Assessment

Gait parameters were acquired using the GAITRite walkway system (CIR Systems Inc., Franklin, NJ, USA). Participants walked barefoot under several conditions: normal walking, i.e. self-selected speed, slow speed, fast speed, head reclined upwards, eyes closed, naming dual task (naming trials: first names, food items, professions, cities), serial subtraction dual task (subtracting 7 from different starting numbers), motor dual task (walking while carrying a tray). Participants were allowed to rest between conditions to minimize fatigue effects. Each condition required participants to walk four lengths (two forward and two backward). Dual-tasking and sensory-challenging conditions place distinct demands on attentional resources, sensorimotor integration, and postural control[27]. Extracted parameters were in line with established gait domains[28], including gait velocity and stride time variability (STV, coefficient of variation of stride time) as a measure gait automaticity (with a higher STV indicating a lower automaticity[29,30]). Each parameter was averaged across valid trials.

### Postural Assessment

Static postural control was evaluated using posturography (Kistler force plate, NeuroPlatform software). Participants underwent a standardized protocol, which includes conditions varying visual, proprioceptive, and vestibular demands: eyes open; eyes closed; head reclination, eyes open; head reclination, eyes closed; foam standing, eyes open; foam standing, eyes closed; foam standing, head reclination, eyes open; foam standing, head reclination, eyes closed; tandem stance (one foot in front of the other), eyes open; tandem stance, eyes closed. Each trial lasted 20–30 seconds, with participants instructed to maintain balance while minimizing extra movement. Safety measures (light touch, hand support, or examiner assistance) were applied if necessary to prevent falls. Center of pressure (CoP) displacement signals were recorded and CoP swaypath in the mediolateral (X) and anteroposterior (Y) directions was extracted. A frequency-domain analysis of CoP, employed to identify altered postural control strategies[31], was performed[26] with spectral power computed separately for the X and Y axes within the following frequency bands: 0–2.4 Hz, 2.4–3.9 Hz, 3.9–8 Hz, and 11–19 Hz (based on[32]).

### Statistical Analysis

Clinical, personality, and neuropsychological data were analyzed using independent samples tests to compare patients with PPPD and healthy controls (HC). Parametric and non-parametric tests were used as appropriate after assumption checks. The Cortisol Awakening Response (CAR) was evaluated by calculating the area under the curve (AUC) with respect to the ground for morning samples[33,34] using Rstudio (Posit, version 2025.09.1). α-amylase values were averaged for each participant. Gait parameters of velocity and automaticity (i.e. STV) were analysed by comparing the baseline condition of normal walking to other conditions, using 2-way repeated-measures Analysis of Variance (rmANOVA). Correlation analysis was performed using Pearson’s correlation. All analyses were two-tailed with a significance threshold of *α* = 0.05 and performed using JASP (Version 0.95.4). Details of statistical analyses are reported in the legends of the corresponding figures and in **Supplementary Methods**.

## Results

### Study participants and demographic characteristics

Out of the 20 recruited PPPD patients and 20 HC, two PPPD patients had to be excluded from further analysis because of insufficient MRI data quality, yielding a final sample of 18 PPPD patients and 20 HC. Although the MRI results are described in another paper, we decided to analyse the same sample of participants also here for comparability. The sample was matched in age and sex (PPPD patients age = 44.8 ± 15.6; HC age = 41.6 ± 15.8, mean ± SD; p=0.588, Mann-Whitney U test; PPPD patients: 9 M – 9 F; HC: 7 M – 13 F; p=0.35, Chi-squared test; intake of psychotropic medication in 7 PPPD patients and in 1 HC; smokers: 1 PPPD patient, 1 HC).

### Self-report Questionnaires

Compared to HC, patients with PPPD reported significantly higher levels of depressive symptoms, but comparable levels of state anxiety, with a trend for increased trait anxiety. Patients also reported higher perceived stress (**Fig.1a**). Regarding health-related quality of life (SF-36), PPPD patients showed reduced *physical functioning* (**Fig.1b**), lower *physical health*, and decreased *social functioning*. Other subdomains did not differ significantly between groups. In their self-report on NQoL, PPPD patients scored lower than HC on all three tested subscales: *ability to participate in social roles and activities, satisfaction with social roles and activities*, and *cognitive function* (**Table 1**). No significant differences were observed between groups on measures of trauma and personality traits (**Fig.1a** and **Table 1**). Assessments of dizziness symptoms and their self-reported and clinician-rated severity showed on average a mild-to-moderate severity (**Fig.1c**). PPPD patients showed a trend for more vertigo-related difficulties overall (NVI), driven by a significant difference in the domain of Vision (**Fig.1d**). Overall, PPPD patients showed increased symptoms of depression and stress, decreased general health and daily hindrance compared to HC, but no differences in past traumatic experiences or personality traits.

**Figure 1.**
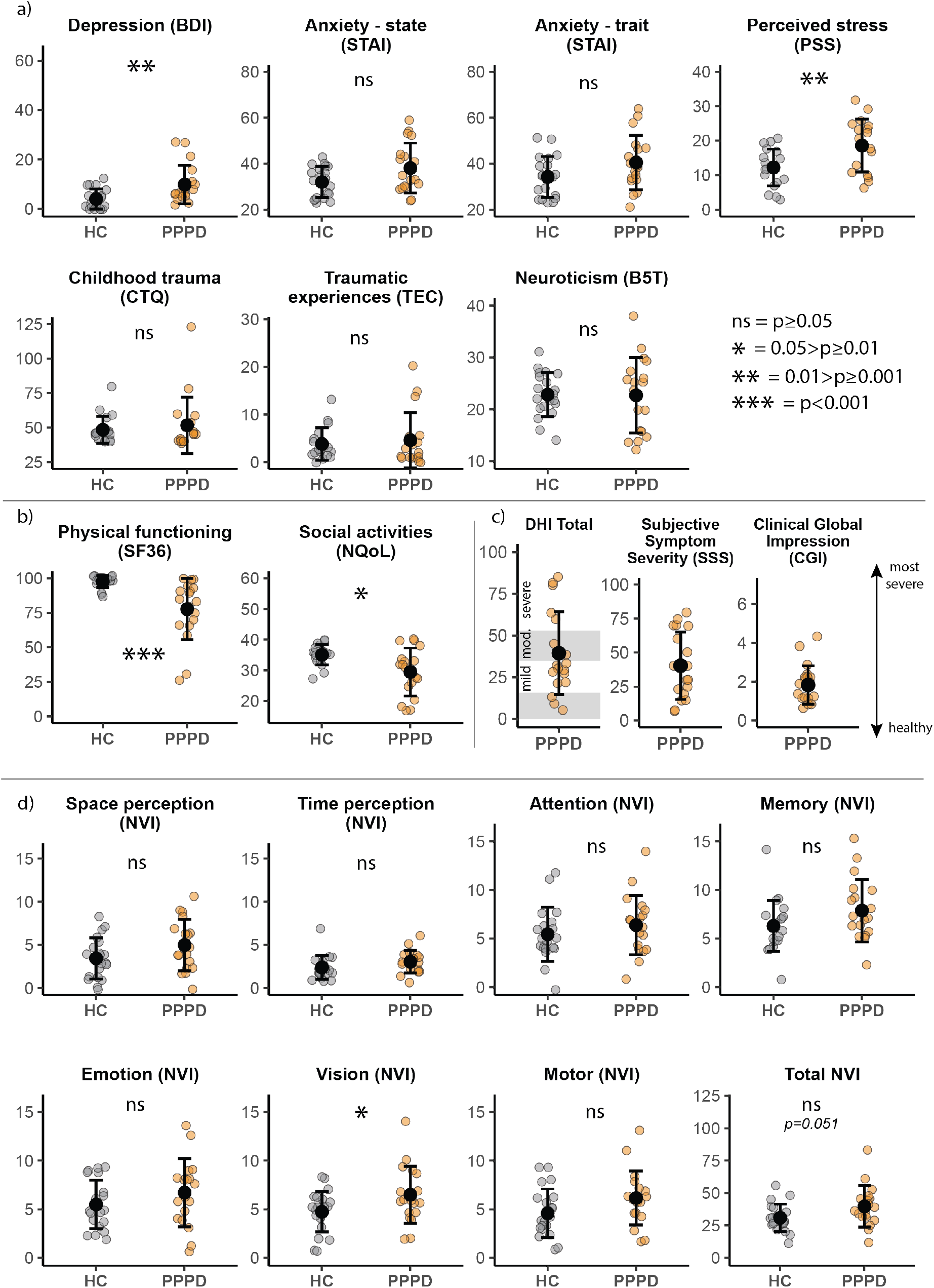
Self-report questionnaires and clinical characteristics of PPPD patients compared to HC. **A)** Self-report questionnaires of mood, stress, trauma and personality traits. PPPD patients report significantly higher depression scores (Beck’s Depression Inventory – BDI: PPPD 9.83± 7.79, HC 4.00 ± 4.03, p=0.004, Mann-Whitney U-test), but not anxiety (State and Trait Anxiety Inventory - STAI: State: PPPD 38.11 ± 10.83, HC 32.05 ± 6.77, p=0.10, Student’s t-test; Trait: PPPD 40.55 ± 11.86, HC: 34.25 ± 8.92, p=0.071, Student’s t-test). They also report significantly higher levels of perceived stress (Perceived Stress Scale – PSS: PPPD 18.611 ± 7.66, HC 12.25 ± 5.33, p=0.005, Student’s t-test). PPPD patients do not report higher levels of childhood trauma (Childhood Trauma Questionnaire - CTQ – Total: PPPD 51.72 ± 20.41, HC 48.50 ± 9.68, p=0.547, Welch’s t-test; Traumatic Experience Checklist - TEC – Total: PPPD 4.61 ± 5.77, HC 3.82 ± 3.43, p=0.625, Student’s t-test). PPPD patients have comparable levels of neuroticism to HC (Big 5 Personality Traits – Neuroticism: PPPD 22.72 ± 7.26 HC 22.85 ± 4.25, p=0.948, Welch’s t-test). **B)** Selected examples of SF36 and NQoL subscales (these and other subscales are further shown in **Table 1**) highlighting the impact of PPPD. PPPD patients report a significant decrease in their physical health (Short-form 36 – SF36 – Physical functioning: PPPD 77.77 ± 22.24, HC 97.75 ±4.43, p<0.001) and a decrease in their ability to take part in social roles and activities (Neurological Quality of Life – NQoL – Ability to participate in social roles and activities: PPPD 29.50 ± 7.82, HC 35.10 ± 3.26, p=0.010, Welch’s t-test) **C)** Clinical assessments. PPPD patients are on average moderately affected by their dizziness in their daily life (Dizziness Handicap Inventory - DHI – Total: 39.56 ± 24.81; details on subscales are shown in **Table 1**). They also report a moderate overall severity of symptoms, both self-rated (Subjective Symptom Severity score - SSS: 40.44 ± 24.85) and clinician-rated (Clinical Global Impression – CGI: 1.83 ± 0.95) **D)** PPPD patients show a trend for a higher total score of the Neuropsychological Vertigo Inventory (NVI), driven by a significantly higher score on the vision subscale, but not other subscales (Total: PPPD 39.78 ± 15.89, HC 31.00 ± 10.61, p=0.051, Student’s t-test; Space Perception p=0.085, Time Perception p=0.143, Attention p=0.328, Memory p=0.103, Emotion p=0.231, Vision PPPD 6.50 ± 2.93, HC 4.75 ± 2.07, p=0.039, Motor p=0.075). (Descriptive statistics are reported as mean ± standard deviation.)

**Table 1.** Further self-report questionnaires on health, traumatic experiences, and personality traits. PPPD patients report significantly lower scores, compared to HC, on the *Physical Functioning, Physical Health*, and *Social Functioning* subscales of the SF36 questionnaire, but not on other subscales (*Emotional Wellbeing, Mental health, Energy/fatigue, Pain and General Health*). They also report significantly lower scores on all the subscales of the Neurological Quality of Life assessment that were administered, related to their participation in social activities and to cognitive function. On the other hand, PPPD patients do not report any significantly different aspect of past traumatic experiences on the Childhood Trauma Questionnaire and Traumatic Experiences Checklist, nor on the Big 5 Personality Traits (See table for detailed statistics; M ± SD = mean ± standard deviation).

| Questionnaire |  | PPPD<br>(M±SD) | HC<br>(M±SD) | df | Statistic | p |
| --- | --- | --- | --- | --- | --- | --- |
| **SF36** |  |  |  |  |  |  |
|  | *Physical functioning* | 77.77 ± 22.24 | 97.75 ± 4.43 | - | 307.50 † | <.001 |
|  | *Emotional wellbeing* | 68.51 ± 44.97 | 93.33 ± 17.43 | - | 229.50 † | 0.064 |
|  | *Physical health* | 55.55 ± 45.82 | 97.50 ± 7.69 | - | 271.00 † | <.001 |
|  | *Mental health* | 35.77 ± 7.57 | 37.40 ± 5.54 | 30.93 | 0.746 ‡ | 0.461 |
|  | *Energy/fatigue* | 35.78 ± 7.57 | 51.50 ± 5.87 | 25.56 | 0.522 ‡ | 0.606 |
|  | *Social Functioning* | 56.94 ± 26.51 | 94.37 ± 11.09 | - | 328.00 † | <.001 |
|  | *Pain* | 76.39 ± 28.75 | 88.87 ± 14.72 | - | 212.00 † | 0.328 |
|  | *General health* | 59.72 ± 11.82 | 58.25 ± 12.28 | 35.82 | -0.376 ‡ | 0.709 |
| **Neurological Quality of Life (NQoL)** |  |  |  |  |  |  |
|  | *Ability to participate in social roles and activities* | 29.50 ± 7.82 | 35.10 ± 3.26 | 22.25 | 2.83 ‡ | **0.010** |
|  | *Satisfaction with social roles and activities* | 26.27 ± 8.17 | 34.80 ± 4.16 | - | 296.00 † | <.001 |
|  | *Cognitive function* | 28.5 ± 6.71 | 33.15 ± 2.27 | 24.031 | 2.67 ‡ | **0.013** |
| **Dizziness Handicap Inventory (DHI) - Total** |  | 39.56 ± 24.81 | - | - | n.a. | n.a. |
|  | *Physical* | 11.89 ± 7.24 | - | - | n.a. | n.a. |
|  | *Emotional* | 13.11 ± 8.57 | - | - | n.a. | n.a. |
|  | *Functional* | 14.56 ± 10.89 | - | - | n.a. | n.a. |
| **Childhood Trauma Questionnaire (CTQ) - Total** |  | 51.72 ± 20.41 | 48.50 ± 9.68 | 23.71 | -0.61 ‡ | 0.547 |
|  | *Emotional abuse* | 9.33 ± 5.50 | 8.85 ± 3.30 | 27.23 | -0.32 ‡ | 0.748 |
|  | *Physical abuse* | 6.67 ± 4.60 | 6.00 ± 2.39 | 24.59 | -0.542 ‡ | 0.593 |
|  | *Sexual abuse* | 6.67 ± 4.70 | 5.85 ± 2.21 | 22.80 | -0.723 ‡ | 0.477 |
|  | *Emotional neglect* | 10.44 ± 5.96 | 11.65 ± 4.69 | 36.00 | 0.696 | 0.491 |
|  | *Physical neglect* | 7.39 ± 3.24 | 6.80 ± 1.73 | 25.40 | -0.688 ‡ | 0.498 |
|  | *Minimization* | 11.11 ± 2.72 | 9.35 ± 3.01 | 36.00 | -1.883 | 0.068 |
| **Traumatic Experience Checklist (TEC<sup>1</sup>) - Total** |  | 4.61 ± 5.77 | 3.82 ± 3.43 | 27.95 | -0.494 ‡ | 0.625 |
|  | *Emotional neglect* | 0.72 ± 1.07 | 0.53 ± 0.87 | 32.32 | -0.584 ‡ | 0.563 |
|  | *Emotional abuse* | 0.83 ± 1.09 | 0.76 ± 0.75 | 30.18 | -0.217 ‡ | 0.830 |
|  | *Physical abuse* | 0.44 ± 0.87 | 0.35 ± 0.61 | 30.67 | -0.367 ‡ | 0.716 |
|  | *Threat* | 0.61 ± 0.85 | 0.35 ± 0.61 | 30.77 | -1.039 ‡ | 0.307 |
|  | *Sexual harassment* | 0.28 ± 0.57 | 0.29 ± 0.59 | 32.78 | 0.083 ‡ | 0.934 |
|  | *Sexual abuse* | 0.22 ± 0.55 | 0.18 ± 0.39 | 30.83 | -0.285 ‡ | 0.778 |
| **Big 5 Personality Traits (B5T)** |  |  |  |  |  |  |
|  | *Neuroticism* | 22.72 ± 7.26 | 22.85 ± 4.25 | 26.809 | 0.065 ‡ | 0.948 |
|  | *Extraversion* | 25.66 ± 4.61 | 26.50 ± 5.37 | 36.00 | 0.067 | 0.613 |
|  | *Conscientiousness* | 27.44 ± 4.35 | 25.90 ± 4.54 | 36.00 | -1.067 | 0.293 |
|  | *Openness to experience* | 27.67 ± 5.19 | 29.05 ± 4.37 | 36.00 | 0.892 | 0.379 |
|  | *Agreeableness* | 33.72 ± 3.95 | 32.70 ± 3.81 | 36 | -0.811 | 0.423 |

‡ Welch's t-test; † Mann-Whitney U test; n.a. = non applicable; <sup>1</sup> = missing data for 3 HC.

### Neuropsychological Assessments

No significant between-group differences were detected on measures of attentional performance. Color-Word interference (Stroop) task performance did not differ significantly across groups. Similarly, working memory as assessed by the Digit Span Test did not show group differences. The Block Tapping Test in the backward condition showed a reduced performance in the PPPD group, only at the uncorrected level but not when correcting for multiple comparisons. A complete overview is provided in **Table 2**. Altogether, PPPD patients did not show cognitive impairments in attention, executive function, and working memory.

**Table 2.** Neuropsychological assessments of attention, executive function, and working memory. PPPD patients show comparable performance to HC on the Test of Attentional Performance (TAP), the Stroop Color-Word Interference Test (Stroop), the Block Tapping test (BTT), and the Digit Span test. They show a significantly reduced performance on the Backward instrument of the BTT, only at the uncorrected level. (See table for detailed statistics; M ± SD = mean ± standard deviation).

| Neuropsychological assessment |  | PPPD<br>(M±SD) | HC<br>(M±SD) | df | Statistic | P<br>(uncorrected) | P<br>(FDR-corrected) |
| --- | --- | --- | --- | --- | --- | --- | --- |
| **Test of Attentional Performance (TAP)** |  |  |  |  |  |  |  |
|  | *Phasic Alertness Reaction Time (RT)* | 46.53 ± 11.91 | 43.55 ± 6.79 | 35.00 | -0.95 | 0.348 | 0.859 |
|  | *Working Memory – Errors (RT)* | 47.00 ± 11.71 | 46.17 ± 11.44 | 24.49 | -0.18 ‡ | 0.859 | 0.859 |
|  | *Working Memory – Omissions (RT)* | 50.94 ± 13.54 | 52.15 ± 9.16 | 35.00 | 0.32 | 0.749 | 0.859 |
| **Stroop Color–Word Interference Test<sup>1</sup>** |  |  |  |  |  |  |  |
|  | *Color naming time* | 29.11 ± 5.03 | 27.84 ± 5.45 | 35.00 | -0.73 | 0.467 | 0.527 |
|  | *Word reading time* | 20.50 ± 3.73 | 18.63 ± 3.73 | 34.89 | -1.52 ‡ | 0.137 | 0.320 |
|  | *Non-corrected errors - Inhibition* | 0.22 ± 0.73 | 0 ± 0 | n.a. | n.a. | n.a. | n.a. |
|  | *Self-corrected errors - Inhibition* | 0.78 ± 1.26 | 1.05 ± 1.35 | 34.99 | 0.64 ‡ | 0.527 | 0.527 |
|  | *Time Inhibition* | 49.55 ± 9.31 | 43.63 ± 10.17 | 35.00 | -1.22 | 0.230 | 0.323 |
|  | *Non-corrected errors - Switching* | 0.39 ± 0.50 | 0.16 ± 0.37 | 31.42 | -1.58 ‡ | 0.124 | 0.320 |
|  | *Self-corrected errors - Switching* | 0.67 ± 0.84 | 0.79 ± 0.85 | 34.95 | 0.44 ‡ | 0.120 | 0.320 |
|  | *Time Switching* | 55.22 ± 10.81 | 50.31 ± 13.57 | 34.03 | -1.22 ‡ | 0.231 | 0.323 |
| **Block Tapping Test (BTT) - Total** |  | 18.44 ± 2.85 | 19.85 ± 3.20 | 36.00 | 1.42 | 0.164 | - |
|  | *Forward* | 9.78 ± 1.63 | 10.00 ± 2.02 | 35.58 | 0.374 ‡ | 0.710 | 0.710 |
|  | *Backward* | 8.61 ± 1.81 | 9.85 ± 1.69 | 36.00 | 2.17 | **0.036** | 0.072 |
| **Digit Span Test** |  |  |  |  |  |  |  |
|  | *Forward* | 7.72 ± 2.24 | 8.15 ± 2.25 | 36.00 | 0.58 | 0.562 | 0.562 |
|  | *Backward* | 6.89 ± 1.57 | 8.15 ± 2.37 | 36.00 | 1.91 ‡ | 0.059 | 0.118 |

‡ Welch's t-test; † Mann–Whitney U test; <sup>1</sup> = one outlier removed; RT = reaction time.
Note: P-values were adjusted for multiple comparisons using the Benjamini–Hochberg FDR procedure within each test (NVI: 8 comparisons, TAP: 3 comparisons, Stroop: 8 comparisons, BTT: 3 comparisons, Digit Span: 2 comparisons).

### Salivary Stress Markers

No significant differences were found between PPPD patients and HC regarding salivary stress markers cortisol and α-amylase (**Fig.2**).

**Figure 2.**
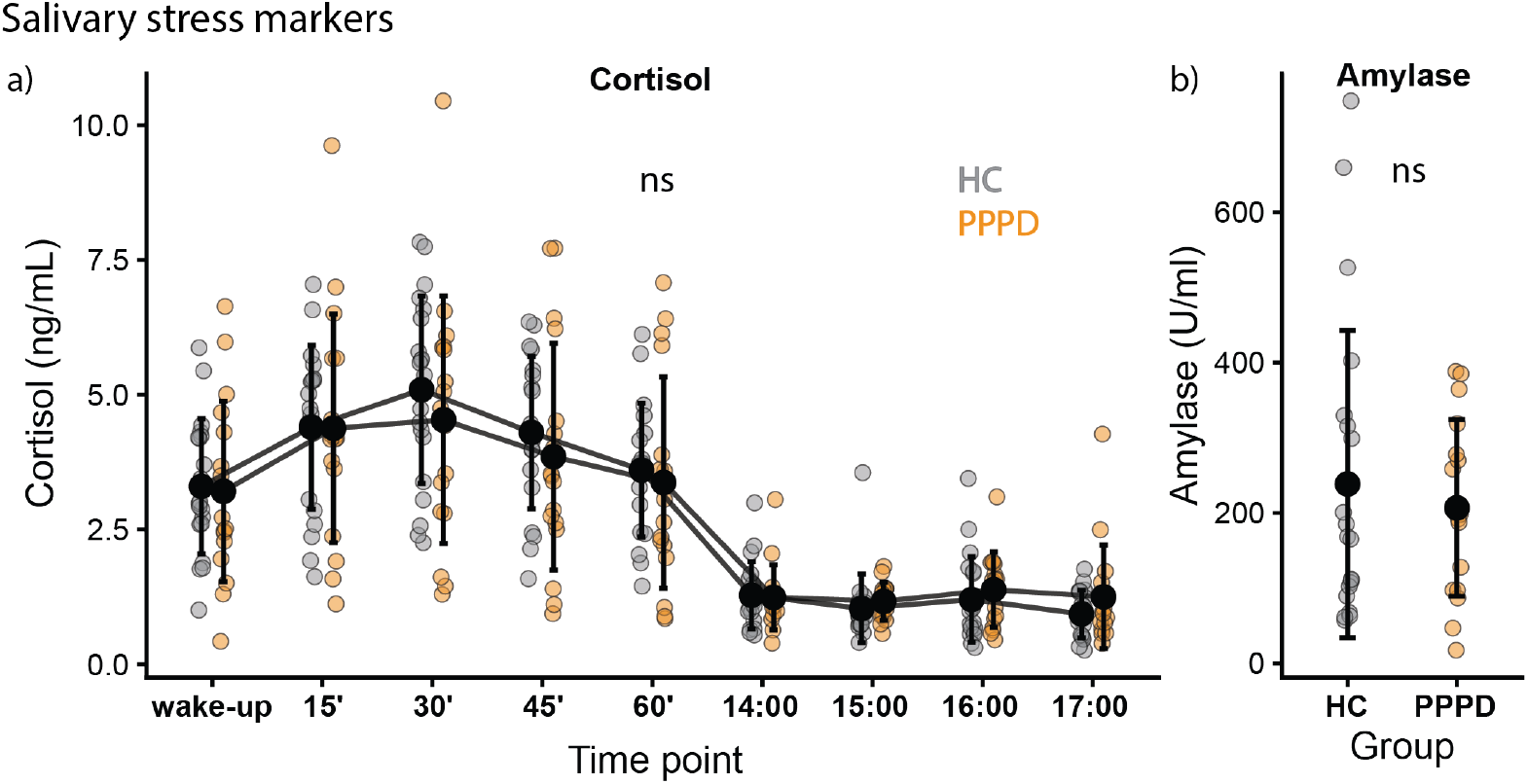
Salivary stress markers (cortisol and α-amylase). PPPD patients show comparable levels of cortisol throughout the day and α-amylase compared to HC. **a)** Levels of salivary cortisol do not differ significantly between PPPD patients and HC. Cortisol samples were collected at different time points throughout the day to capture the Cortisol Awakening Response (CAR) in the morning as well as Diurnal Basal Cortisol (DBC) levels in the afternoon, showing no group effects (the whole-day time course of cortisol was analyzed with a linear model with smoke as a confounding variable: Cortisol ~ timepoint*group + smoke, main effect of group p=0.64, main effect of timepoint p<0.001, main effect of smoke p=0.76, group*timepoint interaction p=0.90; additionally, for the CAR, the area under the curve for morning samples was compared between groups, accounting for smoke as a confounding variable in a linear model: Cortisol ~ group + smoke, main effect of group p= 0.61, main effect of smoke p=0.82). **b)** Salivary α-amylase levels did not differ significantly between PPPD patients and HC, accounting for smoke as a confounding variable (average of 2 samples collected before and after MRI scanning on site; Amylase ~ group + smoke, main effect of group p=0.51, main effect of smoke p=0.01). Black dots represent the mean, error bars represent the standard deviation.

### Gait Assessment

PPPD patients walk generally slower across all speeds, with no further effects of the speed itself (i.e. no group x condition interaction, **Fig.3a**). When asked to walk with their eyes closed, instead, PPPD patients decreased their walking speed significantly more than HC (**Fig.3b**), revealing an increased reliance on vision. When asked to walk with their head reclined backwards or to perform simultaneously a dual task (motor or cognitive), the walking speed of PPPD patients did not deteriorate further (**Fig.3c-d**). STV did not reveal any group differences across walking speeds or other conditions, except for a group effect for head reclination, and the various conditions affected STV similarly in the two groups (**Suppl. Fig.1a-d**). In summary, PPPD patients exhibited a slower gait compared to HC, an increased reliance on vision, but no further differences when subjected to other challenging conditions, and a comparable gait automaticity to HC.

**Figure 3.**
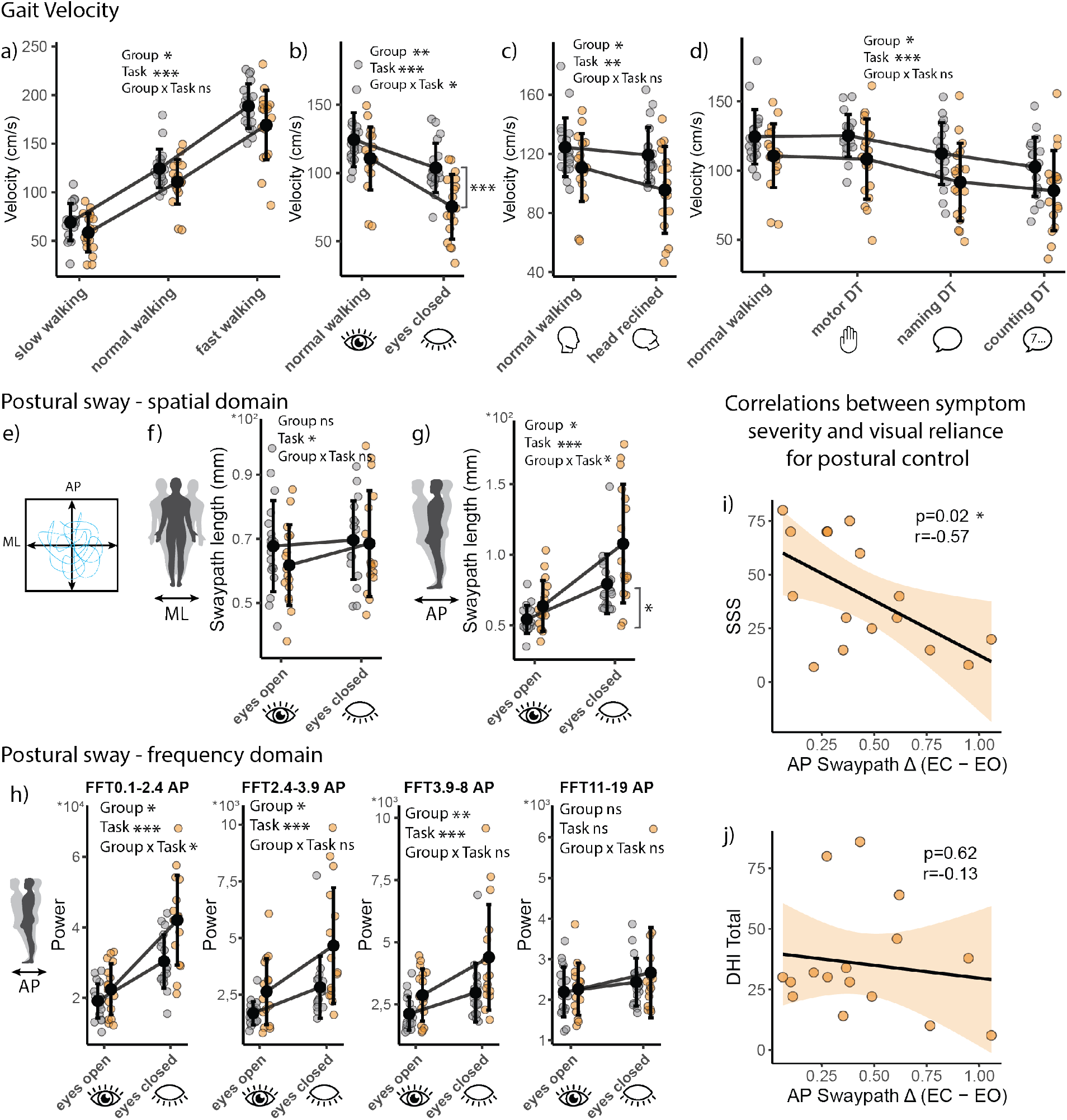
Velocity of gait and postural sway under different conditions in PPPD patients and HC. ***a-d) Gait velocity*. a)** PPPD patients walk generally slower compared to HC when asked to walk at different speeds (rmANOVA, main effect of group p=0.02, main effect of task p<0.001, group x task interaction effect p=0.60). **b)** Walking with eyes closed further decreases the velocity of gait in PPPD patients more than in HC (rmANOVA, main effect of group p<0.01, main effect of task p<0.001, group x task interaction effect p=0.047, post-hoc tests: PPPD normal walking vs HC normal walking p_Holm_=0.14, PPPD eyes closed vs HC eyes closed p_Holm_<0.001, Cohen’s d=1.13). **c)** Walking with their head reclined backwards affects the velocity of gait similarly for PPPD patients and HC (rmANOVA, main effect of group p=0.13, main effect of task p<0.01, group x task interaction effect p=0.13). **d)** Walking while performing motor or cognitive dual tasks affects the velocity of gait similarly for PPPD patients and HC (rmANOVA, main effect of group p=0.02, main effect of task p<0.01, group x task interaction effect p=0.68). Black dots represent the mean, error bars represent the standard deviation. ***e-h) Postural sway*. e)** Illustration of the swaypath (path of motion of the center of pressure, a measure of postural sway) on the posturography board in the anteroposterior (AP) and mediolateral (ML) directions. **f)** Postural sway in the mediolateral (ML) direction is similar in PPPD patients and HC and not affected by the absence of vision (rmANOVA, main effect of group p=0.44, main effect of task p=0.01, group x task interaction effect p=0.14). **g)** Postural sway in the anteroposterior (AP) direction is higher in PPPD patients compared to HC, and exacerbated by the absence of vision (rmANOVA, main effect of group p=0.02, main effect of task p<0.001, group x task interaction effect p=0.02, PPPD eyes open vs HC eyes open p_Holm_=0.13, PPPD eyes closed vs HC eyes closed p_Holm_=0.049, Cohen’s d=-1.13). **h)** Frequency analysis of the postural sway in the AP direction further shows higher power of sway for PPPD patients in frequencies up to 8 Hz, and a trend towards exacerbation by absence of vision in the lowest band (0.1-2.4 Hz: rmANOVA, main effect of group p=0.036, main effect of task p<0.001, group x task interaction effect p=0.04, post-hoc tests non-significant. 2.4-3.9 Hz: rmANOVA, main effect of group p=0.01, main effect of task p <0.001, group x task interaction effect p=0.08. 3.9-8 Hz: rmANOVa, main effect of group p<0.01, main effect of task p= <0.001, group x task interaction effect p=0.12. 11-19 Hz: rmANOVa, main effect of group p=0.39, main effect of task p=0.054, group x task interaction effect p=0.57). Black dots represent the mean, error bars represent the standard deviation. **i-j) *Correlations between visual reliance for postural control and dizziness symptoms*. i)** In PPPD patients, visual reliance for postural control in the anteroposterior (AP) direction (i.e. swaypath difference (Δ) with and without vision) correlates negatively with self-reported symptom severity (p=0.02, r=-0.57). **j)** In PPPD patients, visual reliance for postural control in the AP direction does not correlate with the impact of dizziness on daily life (DHI, p=0.62, r=-0.13). (Pearson’s correlations were performed after removal of one outlier exceeding a swaypath difference above 3*MAD, i.e. median absolute deviation).

### Postural Assessment

Analysis of CoP oscillations in the spatial domain (**Fig.3e**) revealed that PPPD patients did not show significant differences compared to HC in the mediolateral (ML or X) direction (**Fig3f**), but they showed an increase in swaypath length in the anteroposterior (AP or Y) direction, indicative of higher postural instability, exacerbated by the absence of vision (**Fig 3g**). In contrast, other conditions that pose further challenges on postural control did not show a further deterioration of postural stability in PPPD patients compared to HC, neither in the mediolateral nor in the anteroposterior direction (**Suppl. Fig.1e-f**). Analysis of postural sway in the frequency domain for the anteroposterior direction revealed group differences in frequency bands from 0.1 Hz up to 8 Hz. Especially at lower frequencies, closing their eyes affected PPPD patients differently than HC, with a significantly increased power of oscillations in the 0.1-2.4 Hz band (**Fig. 3h**). In contrast, frequency analysis in the mediolateral direction did not show any group differences (**Suppl. Fig.1g**). Overall, the posturographic assessment showed an increased postural instability for PPPD patients in the anteroposterior direction, exacerbated by the absence of vision, especially at low oscillation frequencies.

Since postural instability and reliance on vision are characteristic features of PPPD[6,22], we subsequently calculated the difference between swaypath length in the anteroposterior direction between the eyes closed and eyes open conditions, as an index of reliance on vision for postural control (Δ EC-EO). This measure was negatively correlated with the Subjective Symptom Severity score (**Fig.3i**), but not with other measures of symptom severity (DHI, **Fig.3j**), indicating that a higher reliance on vision is linked to a lower self-perception of dizziness severity.

## Discussion

The present multilevel investigation of PPPD patients and matched HC offers insights into the roles of the different factors known or hypothesized to contribute to the pathophysiology as risk or perpetuating factors.

Clinically, the patients in our sample reported greater depressive symptoms, a trend for increased state anxiety, and higher perceived stress compared to HC. This aligns with previous reports in the literature on PPPD patients[3], as well as in the broader literature on FNDs[2]. PPPD patients also reported a decrease in physical health and a reduced participation in social activities, underscoring the impact of the disorder on everyday life, with an overall moderate severity, comparable to previous reports[4]. Contrary to our hypothesis, past traumatic experiences were not more present in PPPD patients than in HC (or not detectable by the instruments employed here), neither was the personality trait of neuroticism, suggesting that these factors play a lesser role than previously thought. Personality traits have been repeatedly linked to mental health outcomes, however, the meaning of this association has been put into question, as neuroticism might simply reflect the momentary amount of distress[35]

The performance of PPPD patients in neuropsychological tests of attention and executive function revealed no impairment, contrary to our hypothesis. The only lower score observed was at the uncorrected level in the BTT-backward task condition – a working memory task with a motor component. Taken together with the absence of differences in the similar, but verbal-based Digit Span-backward task, this result might be due to motor coordination difficulties rather than cognitive difficulties. Overall, these findings indicate that PPPD is not characterized by global cognitive or attentional impairments *per se*, but they do not exclude that an atypical deployment of attentional happens in relation to postural control. Moreover, similarly to other FNDs[36], the cognitive difficulties self-reported by patients were higher than those observed upon testing.

The levels of salivary stress markers did not show group differences, contrary to what initially hypothesized. This suggests that biological stress responses might play a lesser role in PPPD compared to other FNDs. This contrasts with the levels of perceived stress self-reported by PPPD patients here, an incongruence between subjective and marker-based measures of stress, common in FNDs[19].

In our sample, PPPD patients showed an overall slower gait, which can reflect cautious walking, exacerbated in the absence of vision. This is consistent with previous reports[5,6]. Other conditions where vision is preserved but other systems are challenged did not further decrease gait speed, again underscoring the reliance on vision. This differs from functional gait disorders, where the gait speed is instead affected by dual tasking and the automaticity of gait is altered[37]. These results are consistent with a previous study on PPV[38] and highlight that gait disturbances are not a primary functional alteration in PPPD, but a consequence of dizziness and altered postural control.

Our posturography results confirm again a marked visual dependence in the anteroposterior direction – with an increase in instability with eyes closed – but not in the mediolateral direction. This is consistent with previous studies in PPPD[24,25]. Body sway in the anteroposterior direction depends mostly on ankle adjustments, thus this result likely reflects ankle stiffness, compatible with previous reports[6]. Our findings that additional sensory challenges did not lead to a further deterioration of postural control are consistent with previous findings in PPV[39]. The absence of group differences with dual tasking echoes the distractibility effects seen previously in PPPD[39] and in FND[2], highlighting the role of excessive explicit attention in symptom maintenance.

Our postural frequency analysis shows an increased representation of oscillations in PPPD patients, with a further increase in the absence of vision in the lowest frequency band (0.1-2.4 Hz). This contrasts partially with a previous study on PPV, where an increased sway with eyes closed was found in patients at higher frequencies (3.5-8 Hz), with no differences in swaypath lengths[32]. This discrepancy could be due to differences in the diagnostic criteria of PPV and PPPD[1], and/or to limited sample sizes[32]. Lower oscillation frequencies (0–2 Hz) are considered to reflect visuo-vestibular processes, while higher frequencies (>2 Hz) are considered to reflect proprioceptive processes, thus this result underlines once again the visual dependence for postural control in PPPD patients.

Lastly, a higher reliance on vision was associated with lower subjective severity scores, but not other measures of dizziness-related symptoms. This suggests that visual reliance is employed by patients as a compensatory mechanism and might contribute to the maintenance of PPPD maladaptive postural control strategies by providing the misleading impression of symptom relief.

### Limitations

The sample size employed in this study is limited, however it is in line with previous studies on PPPD[6]. Additionally, our sample differs somewhat from previously reported sex ratios for PPPD, which typically show a higher prevalence in females[40]. Another limitation is that some PPPD patients were medicated with antidepressants and other psychotropic medication, which, along with psychiatric symptoms and comorbidities, might exert an effect on gait and postural sway in either direction. The limited sample size limits the possibility of a subgroup analysis of unmedicated patients.

## Conclusion

This study underlines the impact of PPPD on depressive symptoms, perceived stress, and daily functioning. Additionally, it reveals that past trauma and personality traits do not play a major role in this disorder, thus it does not support a focus on trauma-based interventions. It underscores the central role that visual dependence has in the maintenance and perpetuation of PPPD, both for postural control and gait, thus supporting a therapeutic approach centered on the rebalancing of different inputs for postural control, aimed at reducing the weighting of visual inputs while increasing that of vestibular inputs.

## Data Sharing

The full dataset used for the current study cannot be shared due to ethical and privacy restrictions.

## Authors’ Roles

### CRediT author contributions

Cristina Concetti – *Data Curation, Formal analysis, Project administration, Visualization, Writing – original draft, Writing – review and editing, Conceptualization*; Irene Lozzi – *Data Curation, Formal Analysis, Writing – original draft*; Anna Wyss - *Conceptualization, Methodology, Project administration, Investigation, Data curation;* Salome Häuselmann – *Methodology, Data Curation, Project administration, Investigation*; Tatiana Brémovà-Ertl – *Resources*; Hassen Kerkeni – *Methodology*; Georgios Mantokoudis - *Resources*, Selma Aybek – *Conceptualization, Resources, Supervision, Funding acquisition*.

## Conflicts of Interest

The authors have no conflicts of interest.

## Notes

### Competing Interest Statement

The authors have declared no competing interest.

### Author Declarations

The study was approved by the Ethical Committee of the Canton Bern (SNCTP000004529, 2020-02283), registered on ClinicalTrials.gov (NCT05086380), and conducted according to the principles of the Declaration of Helsinki.

