## Supplementary Materials for "A Multidomain Characterization of Persistent Postural-Perceptual Dizziness Reveals Visual Dependence as Its Main Behavioral Feature"

#### List of abbreviations

AP: Anteroposterior

AUC: Area Under the Curve

BDI-II: Beck Depression Inventory (II)

Big5T: Big Five Personality Test

BTT: Block Tapping Task/Test

CAR: Cortisol Awakening Response

CGI: Clinical Global Impression (scale)

CoP: Center of Pressure

CSD: Chronic Subjective Dizziness

CTQ: Childhood Trauma Questionnaire

DHI: Dizziness Handicap Inventory

F: female

FDR: False Discovery Rate

FFT: Fast Fourier Transform

FND(s): Functional Neurological Disorder(s)

HC(s): Healthy Control(s)

HPA axis: Hypothalamic-Pituitary-Adrenal axis

ICD-11: International Classification of Diseases, 11th revision

M: male

ML: Mediolateral

MRI: Magnetic Resonance Imaging

NQoL: Neurological Quality of Life questionnaire

NVI: Neuropsychological Vertigo Inventory

PPPD: Persistent Postural-Perceptual Dizziness

PPV: Phobic Postural Vertigo

PSS: Perceived Stress Scale

rmANOVA: repeated-measures Analysis of Variance

SAM axis: Sympatho-Adrenomedullar axis

SD: Standard Deviation

SF-36: 36-item Short Form Health Survey

SMD: Space-Motion Discomfort

SSS: Subjective Symptom Severity (score)

STAI (STAI-S, STAI-T): State and Trait Anxiety Inventory (State subscale, Trait subscale)

STV: Stride Time Variability

TAP: Test of Attentional Performance

TEC: Traumatic Experiences Checklist

VV: Visual Vertigo

### Supplementary figures

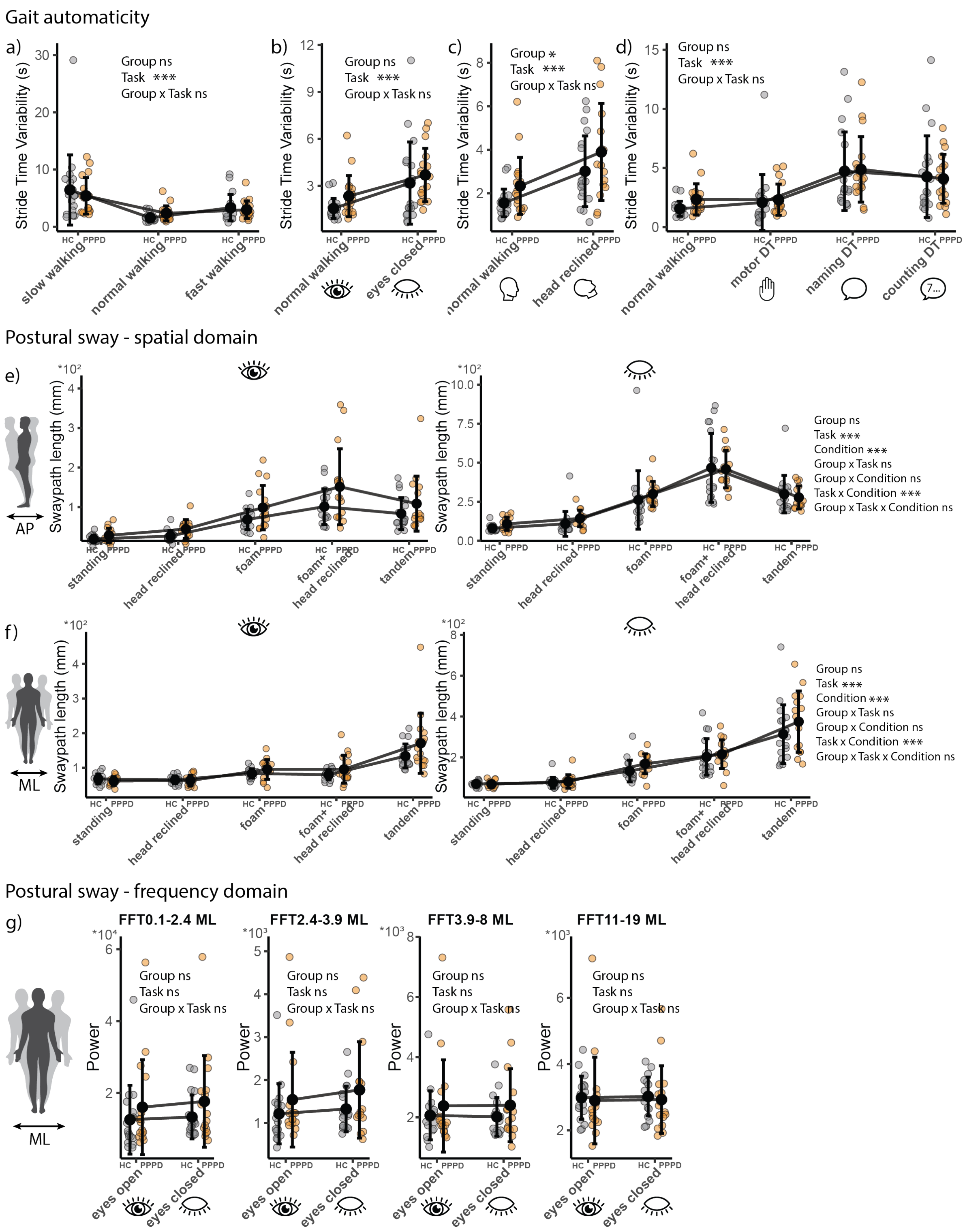

**Supplementary figure 1. Automaticity of gait (stride time variability) and postural sway in the spatial domain under further postural challenges in PPPD patients and HC.** ***a-d) Gait automaticity*.** **a)** Gait automaticity does not differ between groups across different walking speeds (rmANOVA, main effect of group p=0.73, main effect of task p<0.001, group x task interaction effect p=0.46). **b)** Gait automaticity does not differ between groups across presence or absence of vision (rmANOVA, main effect of group p=0.11, main effect of task p<0.001, group x task interaction effect p=0.66). **c)** Gait automaticity is not differently affected by head reclination in PPPD patients and HC (rmANOVA, main effect of group p=0.04, main effect of task p<0.001, group x task interaction effect p=0.99). **d)** Gait automaticity does not differ between groups when performing motor and cognitive dual tasks (rmANOVA, main effect of group p=0.58, main effect of task p<0.001, group x task interaction effect p=0.83). ***e-f) Postural sway in the spatial domain under further postural challenges*.** **e)** Postural sway in the anteroposterior (AP) direction is not differently affected by further postural challenges (condition factor) combined with presence or absence of vision (task factor) in PPPD patients and HC (rmANOVA, main effect of group p=0.24, main effect of task p<0.001, main effect of condition p <0.001, group x task interaction effect p=0.60, task x condition interaction effect p<0.001, group x condition interaction effect p=0.82, group x task x condition interaction p=0.25). **f)** Postural sway in the mediolateral (ML) direction is not differently affected by further postural challenges (condition factor) combined with presence or absence of vision (task factor) in PPPD patients and HC (rmANOVA, main effect of group p=0.16, main effect of task p<0.001, main effect of condition p <0.001, group x task interaction effect p=0.46, task x condition interaction effect p<0.001, group x condition interaction effect p=0.09, group x task x condition interaction p=0.90). **g) *Postural sway in the frequency domain in the mediolateral direction*.** Frequency analysis of the postural sway in the mediolateral (ML) direction shows no differences between PPPD patients and HC (0.1-2.4 Hz: rmANOVA, main effect of group p=0.30, main effect of task p=0.22, group x task interaction effect p=0.21; 2.4-3.9 Hz: rmANOVA, main effect of group p=0.27, main effect of task p=0.19, group x task interaction effect p=0.22; 3.9-8 Hz: rmANOVA, main effect of group p=0.15, main effect of task p=0.39, group x task interaction effect p=0.31; 11-19 Hz: rmANOVA, main effect of group p=0.39, main effect of task p=0.054, group x task interaction effect p=0.57).

### Supplementary Methods

#### Clinical and Personality Questionnaire references

Beck Depression Inventory II (BDI-II[1,2]),

State and Trait Anxiety Inventory (STAI[3]).

36-item Short Form Health Survey (SF-36[4])

Neurological Quality of Life (NQoL[5]).

Perceived Stress Scale (PSS[6]).

Big Five Personality Test (Big5T[7,8]).

Traumatic Experiences Checklist (TEC[9]),

Childhood Trauma Questionnaire (CTQ[10]).

Neuropsychological Vertigo Inventory (NVI[11])

Dizziness Handicap Inventory (DHI[12]).

Clinical Global Impression scale (CGI[13]).

#### Neuropsychological test references

Test of Attentional Performance (TAP[14])

Stroop Color–Word Interference Test[15].

Block Tapping Task (BTT[16]),

Digit Span forward and backward (Wechsler Adult Intelligence Scale[17]).

#### Participant instructions for cortisol sampling

Participants were instructed to follow best practice guidelines[18], including refraining from strenuous physical activities, heavy meals, fruits or fruit juices, coffee, carbonated soft drinks, chewing gum and smoking. Participants were instructed to collect several saliva samples over the course of a single day: immediately after waking up, at 15, 30, 45 and 60 min post-awakening, then at 2, 3, 4, and 5 PM. Participants also noted any delay in sampling time.

#### Statistical analysis

Clinical, personality, and neuropsychological data were analyzed using independent samples tests to compare patients with PPPD and healthy controls (HC). The choice between parametric and non-parametric tests was based on the distribution normality and homogeneity of variance of each variable. Age and sex were not regressed out as our groups were matched for these variables. For the cognitive assessments, p-values are reported both uncorrected and corrected, with FDR correction applied within each test family, as the subscales of each test may tap into overlapping constructs[55].

For salivary stress markers, we evaluated the cortisol curve and the levels of α-amylase. Cortisol samples collected with a delay >5 minutes for morning samples and >15 minutes for afternoon samples were excluded from analysis. The Cortisol Awakening Response (CAR) was evaluated by calculating the area under the curve (AUC) with respect to the ground for morning samples[46,56]. α-amylase values were averaged for each participant. In these analyses, smoke was used as a confounding variable, due to its known influence on salivary markers[19–21]). Age and sex were not used as confounding variables as our groups were matched for these variables and past evidence reports no effects on salivary markers at baseline[22–24]. Similarly, the effects of the menstrual cycle phase[25,26] and menopause[27–29] are very limited or mixed in the literature.

Gait parameters of velocity and automaticity (i.e. stride time variability, STV) were analysed by comparing the baseline condition of normal walking to other conditions, using 2-way repeated-measures Analysis of Variance (rmANOVA). Postural data were analyzed both in the spatial and frequency domains comparing the eyes open and eyes closed conditions using 2-way rmANOVAs. Other conditions that posed further postural challenges were analysed using 3-way rmANOVAs with *task* (two levels: eyes open or closed) and *condition* (five levels: standing, standing with head reclined backward, standing on foam, standing on foam with head reclined backward, tandem standing) as within-subject factors. When significant main effects or interactions involving *group* were observed, post-hoc pairwise comparisons were performed with correction for multiple comparisons.

Correlation analyses with symptom variables were performed only for PPPD patients, after an initial filtering for outlier data points (beyond 3 median absolute deviations), resulting in the exclusion of 1 patient with especially high postural instability. Data normality distribution was checked, and Pearson's correlation was run.
